# Consensus Recommendations for the Clinical Management of Wolfram syndrome Using a Delphi Method

**DOI:** 10.64898/2026.07.02.26357130

**Authors:** Josephine Elliott, Saumel Ahmadi, Patrick Yu-Wai-Man, Sarah Gladstone, Stephanie Gebel, Tracy Lynch, Timothy Barrett, Fumihiko Urano, International Wolfram Syndrome Clinical Guidelines Consortium

## Abstract

**Background:** Wolfram syndrome is a rare neurodegenerative disorder, most commonly caused by pathogenic variants in *WFS1*, while cases due to *CISD2* are exceedingly rare. The estimated prevalence is 1 in 160,000 to 770,000 individuals worldwide. In these clinical guidelines, disorders caused by *WFS1* are referred to as *WFS1*-Wolfram syndrome, and those caused by *CISD2* as *CISD2*-Wolfram syndrome. Historically, it has been characterized by early-onset, antibody-negative, insulin-dependent diabetes mellitus, progressive optic atrophy, sensorineural hearing loss, arginine vasopressin deficiency, and brainstem and cerebellar atrophy. More recently, partial and late onset forms have been identified. There are currently no licensed disease-modifying treatments, and international clinical guidelines have not previously been established.

**Methods:** An international steering committee systematically reviewed 273 peer-reviewed publications and generated draft consensus statements across six clinical domains. These statements were evaluated by international specialists in endocrinology, clinical genetics, neurology, ophthalmology and neuro-ophthalmology, psychiatry, and urology, drawn from North America, Europe, Latin America, Oceania, and Asia, using a modified three-round Delphi process. Additional feedback was incorporated from nurses specializing in multidisciplinary Wolfram syndrome care, from leaders of international patient organizations, and from specialists in the genetic diagnosis of monogenic diabetes. Structured feedback from patients and families was gathered through multiple international patient advocacy organizations. Consensus was defined as ≥80% agreement.

**Results:** All 35 final consensus statements reached the pre-specified consensus threshold of ≥80% agreement, spanning diagnosis and genetic testing, multidisciplinary care organization, neuro-ophthalmology, neurology, endocrinology, urology, gastroenterology, and psychiatry.

**Conclusions:** These guidelines are the first international clinical consensus for Wolfram syndrome and provide actionable recommendations for clinicians worldwide. Implementation should be accompanied by a prospective audit to expand the evidence base and support future iterations.

## Introduction

Wolfram syndrome (OMIM #222300; also known as DIDMOAD: *D*iabetes *I*nsipidus, *D*iabetes *M*ellitus, *O*ptic *A*trophy, *D*eafness) is a rare monogenic, multisystem neurodegenerative disease first described by Wolfram and Wagener in 1938 in four siblings presenting with juvenile-onset diabetes mellitus and progressive optic nerve atrophy^1^. The disease is caused predominantly by biallelic loss-of-function variants in the *WFS1* gene, which encodes Wolframin, a transmembrane glycoprotein of the endoplasmic reticulum (ER) critical for ER calcium homeostasis, the unfolded protein response, and pancreatic β-cell survival^2,3^. A small number of patients have pathogenic variants at a second locus, *CISD2*, which is associated with additional gastrointestinal symptoms and platelet disorders^4^.

Over the past three decades, the clinical phenotype of Wolfram syndrome has expanded considerably. In addition to diabetes mellitus and optic atrophy, individuals develop sensorineural deafness, arginine vasopressin deficiency (diabetes insipidus), progressive brainstem and cerebellar neurodegeneration, cerebellar ataxia, autonomic failure, psychiatric illness (depression and psychosis), and neurogenic bladder^5^. There are no approved disease-modifying treatments. Premature death has been reported at a median age of approximately 30 years, most commonly from brainstem failure and respiratory compromise^5^. However, there are affected people living into their seventh decade^6^. In addition, dominantly inherited, partial phenotypes have also been described^7–9^.

Despite its clinical complexity and global distribution, no international clinical guidelines for Wolfram syndrome have previously existed. Individual clinicians and centers have developed local protocols in isolation, resulting in marked heterogeneity in diagnostic approaches, monitoring schedules, and management practices. Prior to this work, the only consensus documents published for Wolfram syndrome were limited in scope, lacked a formal multi-round methodology, and did not incorporate patient and family perspectives.

Here, we present the first international consensus statement for the diagnosis and clinical management of Wolfram syndrome, developed through a modified three-round Delphi process. The structure and precedent for this work follows the approach established for Cornelia de Lange syndrome, another rare autosomal recessive multisystem genetic disease^10^, Duchenne muscular dystrophy^11^, pituitary adenoma^12^, Beckwith–Wiedemann syndrome^13^, spinal muscular atrophy^14^, and Alström syndrome^15^. These guidelines address the full clinical spectrum of Wolfram syndrome - molecular diagnosis, multidisciplinary care coordination, and organ-specific management - providing actionable recommendations for clinicians worldwide.

## Methods

### Steering committee and systematic review

An international steering committee was established in December 2024, co-chaired by Fumihiko Urano (Washington University, St. Louis, USA) and Timothy Barrett (University of Birmingham, Birmingham, UK). The committee comprised eight members (four in the United States, four in the United Kingdom), including geneticists, an endocrinologist, a neurologist, a neuro-ophthalmologist, pediatricians, and members of two patient advocacy organizations (one from the United States and one from the United Kingdom). All physicians on the steering committee had clinical experience actively managing at least 20 patients with Wolfram syndrome. The committee conducted a structured PubMed review of 273 peer-reviewed English-language articles published from 1996 through early 2026, covering Wolfram syndrome type 1 and type 2, WFS1-related disorders, Wolfram-like syndrome, DIDMOAD, and the WFS1 Spectrum disorder in adults and children. Pre-clinical studies, review articles, editorials, conference abstracts, and non-English articles were excluded. References were collated in Covidence, and three steering committee members reviewed each article for inclusion based on clinical relevance to diagnosis and management. The committee then met every two weeks over six months to draft 41 candidate consensus statements across six clinical domains: diagnosis and general management; neuro-ophthalmology; neurology; endocrinology; urology and gastroenterology; and psychiatry and cognition.

### Expert panel

An international panel of clinicians was recruited to participate in the Delphi process. Panelists were selected on the basis of recognized expertise in Wolfram syndrome or the relevant clinical subspecialty. Specialists were drawn from North America, Europe (United Kingdom, France, Germany, Italy, Denmark, Spain, Belgium, Poland, Switzerland), Georgia, Latin America (Brazil), Oceania (New Zealand), and Asia (Japan, India, Taiwan). The panel spanned the relevant clinical disciplines, including endocrinology, clinical genetics, neurology, ophthalmology and neuro-ophthalmology, psychiatry, urology, pediatrics, and family medicine, together with allied health professionals comprising clinical nurse specialists, clinical psychologists, and a transition coordinator. Panelists’ identities were masked to reviewers during voting rounds.

### Delphi process

A modified three-round Delphi methodology was adopted because it is well-suited to rare diseases such as Wolfram syndrome^16^: it systematically integrates anonymous feedback from international experts and refines candidate statements through iterative steering committee meetings and topic-focused expert sub-group discussions. In each round, panelists rated their level of agreement with each statement on a five-point Likert scale (1 = strongly disagree; 5 = strongly agree) and were able to provide free-text commentary. Panelists were instructed not to rate statements outside their area of clinical expertise. Consensus was defined a priori as agreement (Likert score of 4 or 5) by ≥80% of respondents, in keeping with the upper end of the accepted threshold for Delphi methodology in healthcare. Statements that failed to reach this threshold were revised, merged, or removed based on respondent comments and recirculated in the subsequent round. Up to three rounds of voting were performed; Rounds 1, 2, and 3 received responses from 56, 37, and 32 panelists, respectively. All 35 final consensus statements reached the pre-specified threshold (Figure 1).

**Figure 1.**
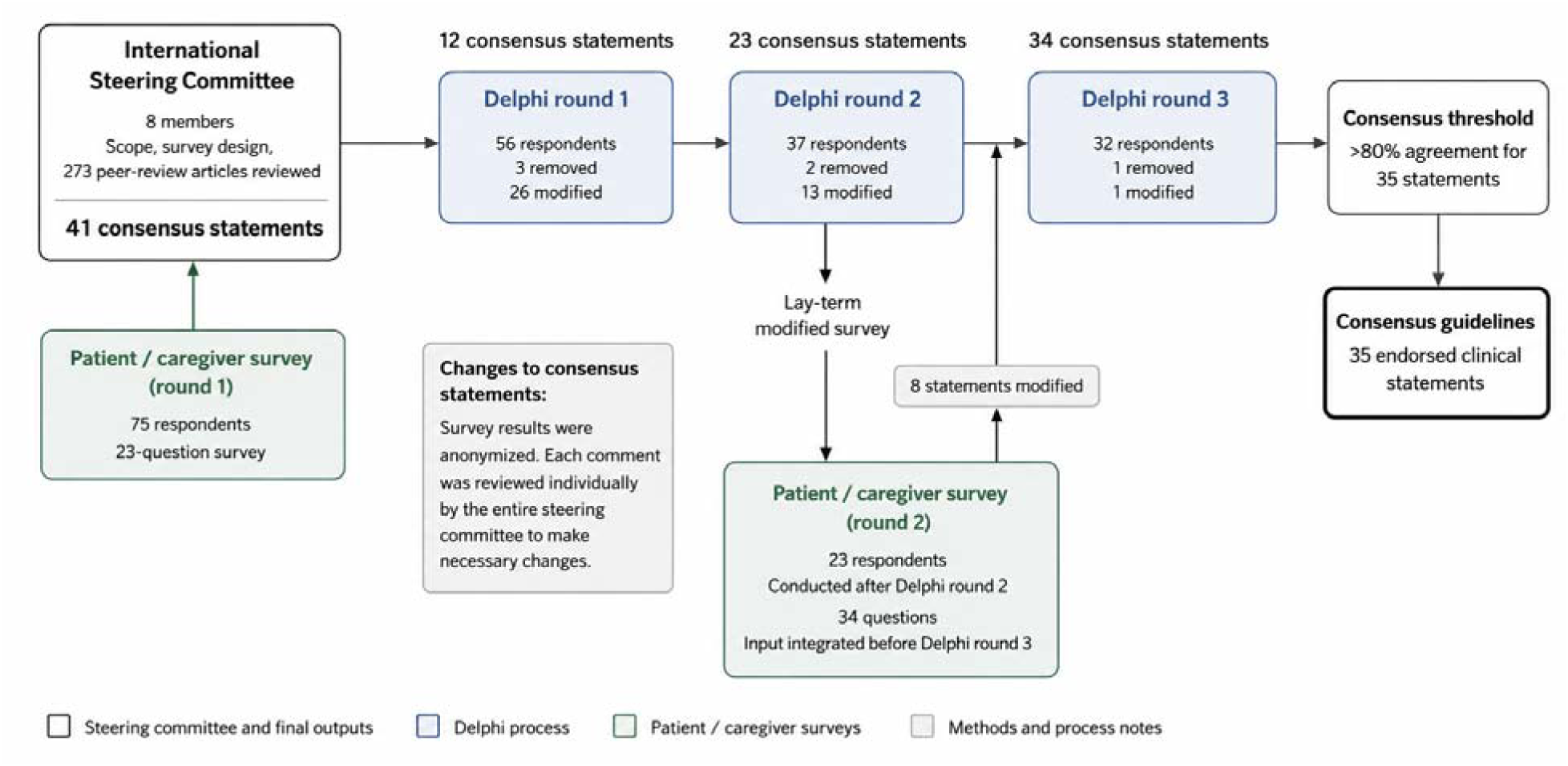
Modified Delphi process for the development of international consensus clinical guidelines for Wolfram syndrome. An international steering committee of 8 members defined the scope and survey design based on a review of 273 peer-reviewed manuscripts and input from patients and caregivers (23-question survey). A three-round Delphi survey using a 5-point Likert scale, and an optional space for comments for each statement, was administered to an international expert panel (Round 1: n = 56; Round 2: n = 37; Round 3: n = 32 respondents). Between rounds, statements were modified or removed based on comments or if they did not reach the pre-defined consensus threshold of 80% (Round 1: 26 modified, 3 removed; Round 2: 13 modified, 2 removed; Round 3: 1 modified, 1 removed). For each revision, the steering committee discussed every free-text comment from survey respondents. A parallel patient/caregiver survey (34 lay-term questions, conducted after Round 2; 23 respondents) was integrated before Round 3. Survey results were anonymized, and each comment from patient/caregiver was also reviewed individually by the steering committee. The entire process yielded 35 endorsed clinical consensus statements with > 80% agreement.

**Figure 2.**
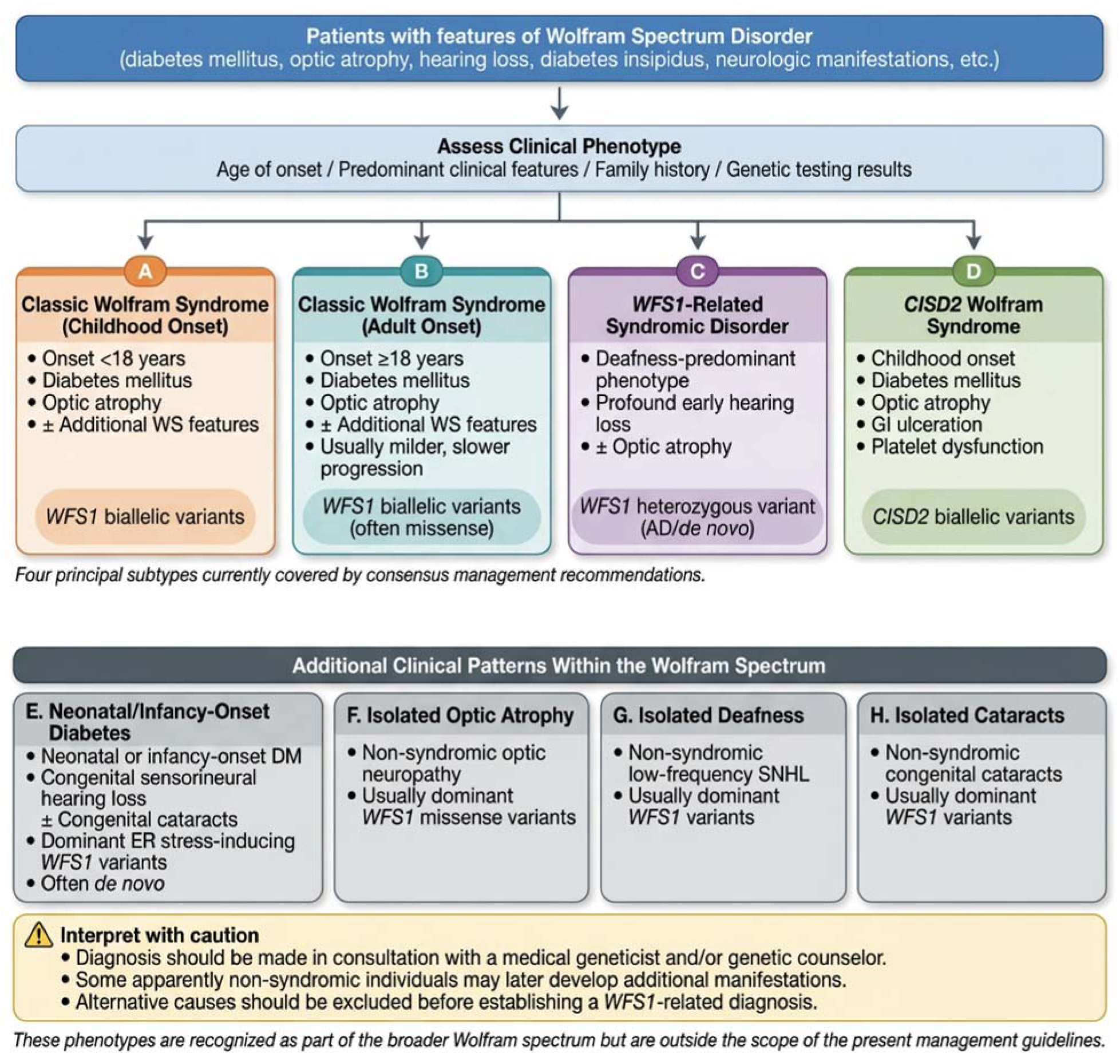
Diagnostic algorithm for Wolfram Spectrum Disorders. Patients are classified into one of four principal Wolfram syndrome subtypes (A–D) based on age of onset, predominant clinical features, and genetic findings. Four additional WFS1-related clinical patterns (E–H), including neonatal/infancy-onset disease and isolated optic atrophy, are shown to represent the full eight-pattern Wolfram spectrum disorder; patterns E–H fall outside the scope of these management guidelines.

### Patient consultation

To ensure that the final guidelines were patient-centered, a parallel patient and family survey was conducted through international advocacy organizations: the Snow Foundation, the Ellie White Foundation, the Unravel Wolfram Syndrome Fund, Be A Tiger Foundation, the Eye Hope Foundation, the Ontario Wolfram League, the Gentian Association, the Alliance of Families Affected by Wolfram Syndrome, the Wolfram Heroes Association, Wolfram Syndrome UK, and the Association du Syndrome de Wolfram. Patients and families are uniquely positioned to identify priorities, lived-experience challenges, and outcomes that matter most in everyday life. Twenty-four lay-language items, thematically aligned with the specialist statements, were rated on the same five-point Likert scale, with free-text commentary invited for each statement. The survey was distributed after Round 2 of the specialist Delphi, allowing candidate statements to first be refined by clinical expert input and then re-examined through the lens of patient and family priorities. Responses were analyzed by the steering committee and used to refine the wording, scope, and emphasis of statements before the final specialist round (Round 3). Patient feedback informed the diagnostic classification, monitoring frequency, and patient-centered management priorities for the most disabling symptoms.

### Evidence grading system

**B** Evidence from well-designed observational studies (cohort or case-control studies), or from systematic reviews of such studies. The body of evidence is sufficient to support the recommendation but is not derived from randomized controlled trials.

**C** Evidence from case series, individual case reports, or expert opinion supported by mechanistic or physiological rationale. The evidence base is limited in quantity or quality.

### Expert opinion

Recommendation based on the formal consensus of the international expert panel in the absence of sufficient published evidence to assign a higher grade. These recommendations reflect current best clinical practice as agreed by specialists with direct experience managing Wolfram syndrome.

### Patient feedback

Recommendation shaped by structured input from patients and families through international advocacy organizations (see Patient consultation section for the full list of participating organizations). Patient perspectives were formally integrated into the Delphi process through parallel rounds of voting.

### Delphi Round

The round in which the statement first reached the pre-specified consensus threshold of ≥80% agreement. Statements reaching consensus in Round 1 were accepted as final without further iteration; those reaching consensus in Rounds 2 or 3 were revised in response to panelist feedback before re-voting.

No statement could be assigned Level A, as there are no randomized controlled trials or meta-analyses of randomized controlled trials in Wolfram syndrome. The absence of randomized controlled trial data to inform clinical practice is itself a key finding of this work and represents an important unmet need.

## Results

The consensus panel comprised specialists from multiple clinical and allied health disciplines who participated in the Delphi process conducted from July 2025 through April 2026.

Participants completed three rounds of voting and were drawn from the clinical specialties and allied health professions described in the Methods.

In Round 1, 41 statements reached consensus. Following review of the panelists’ free-text comments, 26 statements were revised and three were removed, resulting in 12 finalized statements. The 26 revised statements were carried forward to Round 2, where all achieved consensus. Based on additional written feedback, 13 statements were further revised and two were removed, increasing the total number of finalized statements to 23. Following additional feedback from patient surveys, three of these 13 revised statements were modified before Round 3. All 13 revised statements reached consensus in the final round; however, one statement was removed based on panelists’ written comments, resulting in 34 finalized statements. After completion of Round 3, one additional statement was revised in response to panel feedback, yielding a final set of 35 consensus statements (Figure 1).

Similarly designed statements also reached consensus among patient and family respondents. Patient feedback primarily helped shape the definition of Wolfram syndrome, frequency of clinical assessments, and patient-centered management priorities for the most disabling symptoms.

## Recommendations by clinical domains

### Diagnosis and genetic testing

*Statement 1 Wolfram syndrome is caused by biallelic pathogenic variants in WFS1 or CISD2. Diagnosis requires consideration of both clinical pattern and genetic test results, and must be discussed with a medical geneticist and/or genetic counselor. Four clinical subtypes are defined: (A) WFS1-Wolfram syndrome childhood onset; (B) WFS1-Wolfram syndrome adult onset; (C) WFS1-related disorder; (D) CISD2-Wolfram syndrome (consensus* ≥*80%; level of evidence: C / expert opinion; Round 2)*.

The *WFS1* gene on chromosome 4p16 encodes Wolframin, a 100-kDa transmembrane protein with nine transmembrane domains that localizes to the endoplasmic reticulum (ER) and is expressed at highest levels in the brain, pancreatic β cells, and heart^3,17–20^ ^6,21–24^. Biallelic loss-of-function variants cause chronic ER stress, activation of the unfolded protein response, mitochondrial dysfunction, and progressive neuronal and β-cell death^25^. Genotype–phenotype correlations exist: biallelic truncating variants are generally associated with more severe, earlier-onset disease, whereas missense combinations may produce milder phenotypes^6^. The vast majority of patients with Wolfram syndrome carry pathogenic variants in the *WFS1* gene; in these guidelines we refer to this disease as *WFS1*-Wolfram syndrome. The *CISD2* gene on chromosome 4q24 encodes ERIS (NAF-1/miner1), a zinc-finger ER outer-membrane protein involved in iron–sulfur cluster transfer and mitochondrial function^4,26–28^. Biallelic *CISD2* variants cause *CISD2*-Wolfram syndrome with additional gastrointestinal ulceration and platelet defects^4,29^. A spectrum of dominantly inherited *WFS1*-related disorders also exists, ranging from isolated low-frequency sensorineural hearing loss^7^, isolated optic neuropathy^30^, isolated cataracts^31^, combinations of hearing loss and optic neuropathy^8,32,33^ to combinations of diabetes mellitus, optic atrophy, cataracts, hypotonia and developmental delay^9^. These autosomal-dominant *WFS1*-related disorders are related to, but clinically distinct from, autosomal-recessive Wolfram syndrome and are beyond the scope of the present guidelines.

The original description by Dr. Wolfram in 1938 was of four siblings with an inherited association between juvenile-onset diabetes mellitus and progressive optic nerve atrophy^1^. This combination has since become embedded in the medical literature as clinically diagnostic of Wolfram syndrome^5,34^.

Since this original description, it has become apparent that there is a spectrum of conditions associated with pathogenic variants in two genes, *WFS1* and *CISD2*. Within the condition we identify as Wolfram syndrome, there is a wide spectrum of severity. We therefore recommend that clinicians consider both the clinical pattern and the genetic test results in order to make a diagnosis; and that a diagnosis should only be made in discussion with a medical geneticist and/or genetic counselor.

We have categorized conditions caused by variants in *WFS1* and *CISD2* genes into the following sub-categories under the umbrella of Wolfram Spectrum Disorders:

A-*WFS1*-Wolfram syndrome childhood onset

B-*WFS1*-Wolfram syndrome adult onset

C-*WFS1*-related disorder

D-*CISD2*-Wolfram syndrome

Some *WFS1* variants have been associated with specific patterns such as neonatal diabetes, congenital hearing loss, and hypotonia^9^, isolated non-syndromic deafness^7,35^, isolated optic neuropathy^30^, and isolated cataracts^31^. These are associated with specific genotypes and are beyond the scope of this guideline.

A. To make a diagnosis of Wolfram syndrome childhood onset, the patient must have childhood onset (under 18 years) diabetes mellitus and optic atrophy, together with biallelic pathogenic/likely pathogenic variants in the *WFS1* gene (autosomal recessive).

The presence of one pathogenic/likely pathogenic variant in the *WFS1* gene and one variant of unknown significance may confirm a diagnosis of Wolfram syndrome if the clinical picture is consistent with Wolfram syndrome, and these findings must be discussed with a medical geneticist and/or genetic counselor.

B. To make a diagnosis of Wolfram syndrome adult onset, the patient will likely have adult onset (18 years or older) diabetes mellitus and optic atrophy, together with biallelic pathogenic/likely pathogenic variants in the *WFS1* gene (autosomal recessive).

The adult-onset form has been separated from the childhood onset form as it usually has a much slower time-course, and can have milder features. The adult form is usually caused by missense variants in the *WFS1* gene, for instance the variant commonly seen in the Ashkenazi Jewish population^36–38^.

C. To make a diagnosis of *WFS1*-related syndromic disorder, the affected person must have profound early-onset deafness with or without optic atrophy, together with one pathogenic / likely pathogenic variant in the *WFS1* gene occurring either as an autosomal dominant or as a de novo variant.

D. To make a diagnosis of *CISD2* Wolfram syndrome, the patient must have childhood onset (under 18 years) diabetes mellitus and optic atrophy, gastrointestinal symptoms of ulceration and a platelet disorder, together with biallelic pathogenic/likely pathogenic variants in the *CISD2* gene (autosomal recessive) ^4^.

### Disclaimer

Autosomal dominant *WFS1*-related disorders exist along a broad spectrum of presentation. There is a category of *WFS1*-related non-syndromic disorders in which individuals may show isolated findings, such as isolated sensorineural hearing loss, isolated optic neuropathy, or isolated cataracts. Although there is one known dominant *WFS1* variant associated with isolated diabetes, in most cases *WFS1* dominant variants do not cause monogenic diabetes^39,40^. Caution is needed, as some individuals who initially appear to have “non-syndromic” forms may later develop additional features. In addition, patients who seem to have *WFS1*-related syndromic disorders may develop certain symptoms for reasons unrelated to their *WFS1* variants, such as autoimmune type 1 diabetes or antibody-negative type 1 diabetes^41^.

Therefore, for each symptom commonly seen in Wolfram syndrome and *WFS1*-related disorders, other potential causes should be carefully ruled out before concluding that the symptom is *WFS1*-related. The severity, symptoms, and progression vary among individuals. Genetic results should always be interpreted with a qualified health professional, such as a medical geneticist or genetic counselor, as interpretation without appropriate expertise may be misleading.

### Multidisciplinary care organization

*Statement 2 Patients aged under 18 years should be offered an annual multidisciplinary assessment, or sooner if there is significant deterioration. Patients aged 18 years or older should be offered assessment every 1–2 years depending on disease severity, or sooner if there is significant deterioration (consensus* ≥*80%; expert opinion / patient feedback; Round 3)*.

The multisystem nature of Wolfram syndrome necessitates coordinated care from multiple specialties^2,5^. The panel recommends that all patients be assessed by a formal multidisciplinary team (MDT) at regular intervals: annually for patients under 18 years of age and every 1–2 years for adults (depending on disease severity and symptom burden), with earlier review following significant clinical deterioration. The judgment as to what constitutes significant clinical deterioration, and the decision to bring forward a multidisciplinary review, is best made by the patient’s primary or local specialist team, who can coordinate timely assessment without displacing the routine review of other patients.

*Statement 3 A core Wolfram syndrome multidisciplinary team will ideally include a coordinator (geneticist or nurse navigator), endocrinologist, neurologist, ENT/audiology specialist, ophthalmologist or neuro-ophthalmologist with optometry support, and urologist with access to urodynamics. Additional members as clinically indicated may include a genetic counselor, clinical psychologist or psychiatrist, speech and language therapist, physical therapist, gastroenterologist, adult-care transition coordinator, patient support group family liaison, and sleep specialist (consensus* ≥*80%; expert opinion / patient feedback; Round 3)*.

A core Wolfram syndrome MDT should include a coordinator (medical geneticist or nurse navigator), endocrinologist, neurologist, ENT/audiology specialist, ophthalmologist with optometry support, and urologist with access to urodynamic testing. Additional members, including a clinical psychologist/psychiatrist, a genetic counselor, a speech and language therapist, a physical therapist, a gastroenterologist, an adult-care transition coordinator, a patient advocacy liaison, and a sleep specialist, should be included according to individual clinical need and local availability. Given the substantial psychological and neuropsychiatric burden of Wolfram syndrome, access to clinical psychology or psychiatry is particularly important and should be considered a key component of the team wherever possible.

***Statement 4 Upon diagnosis, all patients should be offered contact with a patient advocacy organization and assessment for entitlement to benefits, caregiver support, and (for children or young people) educational support (consensus*** ≥***80%; expert opinion; Round 1)*.**

Patient advocacy organizations are integral partners in care and a valuable source of practical, emotional, and educational support from the time of diagnosis. All patients and families should be signposted to relevant organizations (Table 1) and assessed for entitlement to benefits and caregiver support, with educational support arranged for children and young people.

**Table 1.**
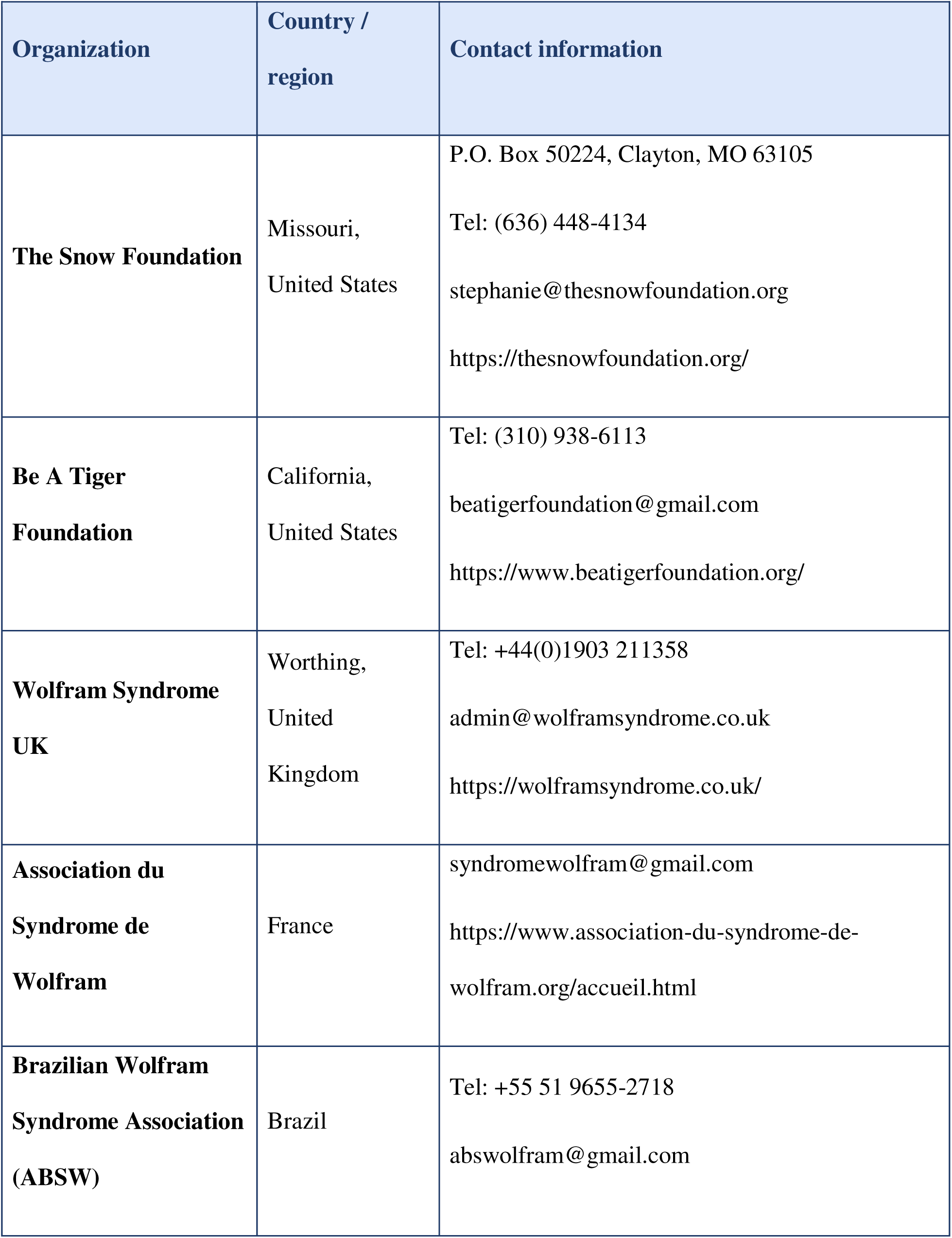
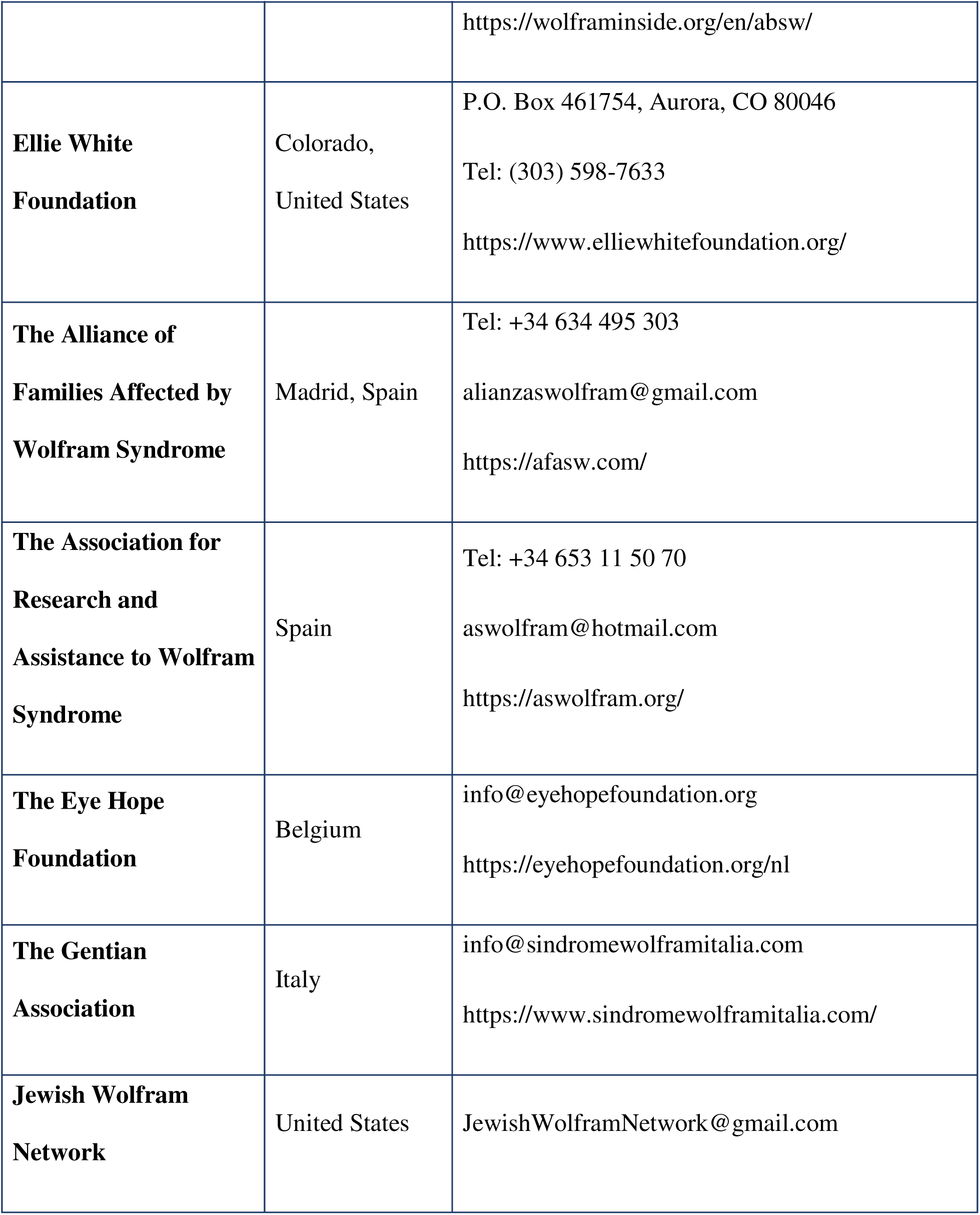
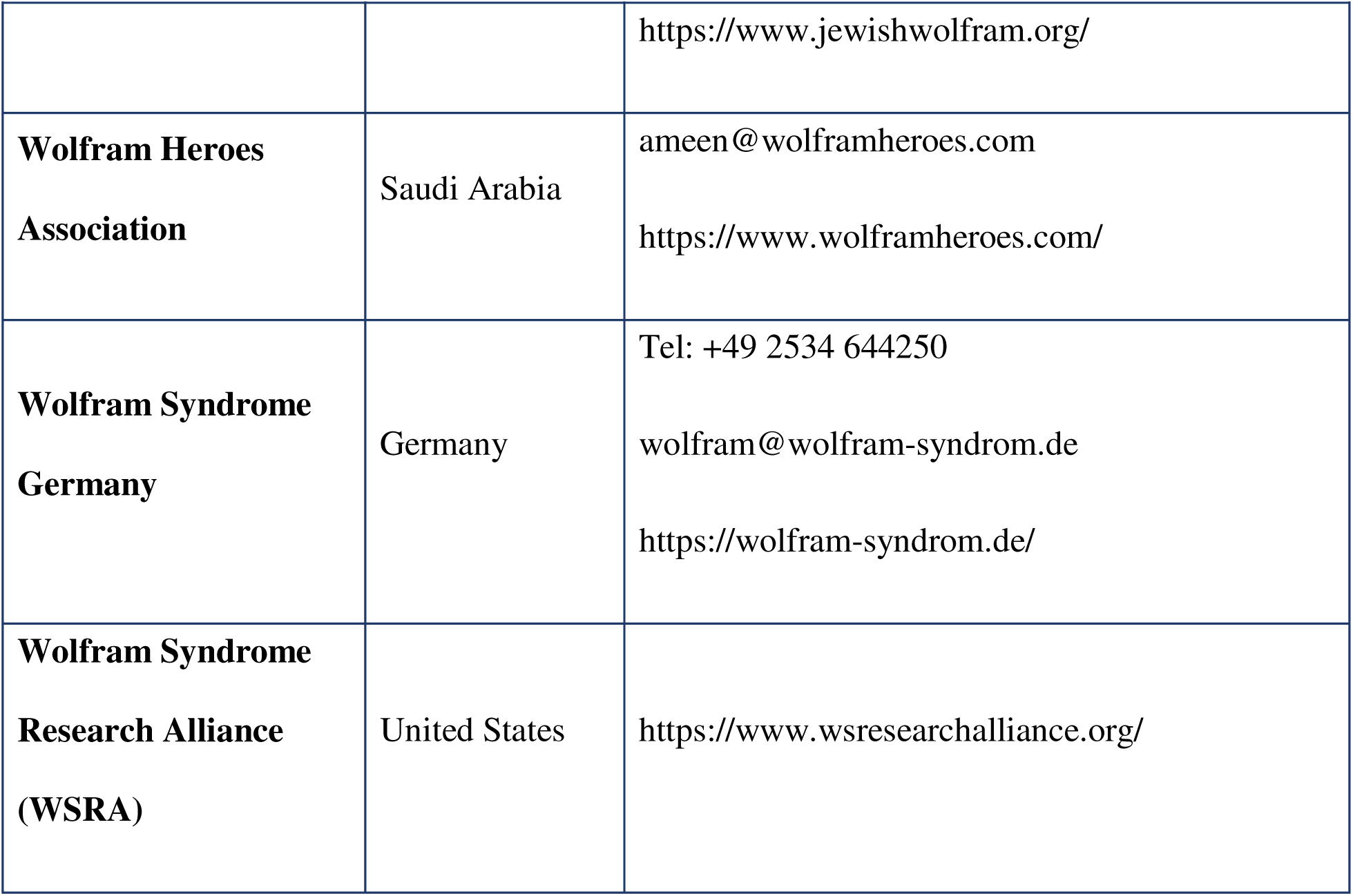
Patient advocacy organizations involved in the development of these guidelines.

***Statement 5 Transition from pediatric to adult care should begin early in the second decade of life. The young person and care providers should both be involved in planned transfer to adult services (consensus* ≥*80%; expert opinion; Round 1)*.**

At diagnosis, all patients and families should be offered contact with patient advocacy organizations and assessment for entitlement to benefits, caregiver support, and educational support. Transition from pediatric to adult care should begin in the second decade of life, with active involvement of the young person in all planning discussions. Use of a structured transition readiness tool, such as the Transition Readiness Assessment Questionnaire^42^ or the Ready Steady Go programme ^43^, can help guide and document this process.

### Neuro-ophthalmology

***Statement 6 Patients with a Wolfram Spectrum Disorder should have an ophthalmological assessment at least once a year, including best-corrected visual acuity (cycloplegic refraction in children), color vision, intraocular pressures, slit-lamp examination of the anterior and posterior segments, visual fields (if age-appropriate), and OCT imaging of the optic nerve and macula (if age-appropriate) (consensus*** ≥***80%; expert opinion; Round 2)*.**

Progressive optic atrophy typically presents in the first or second decade of life and progresses to legal blindness in most patients. Visual loss begins with deficits in color vision and central visual field defects. Optical coherence tomography demonstrates early retinal nerve fiber layer thinning, which can precede visual symptoms.

Progressive optic atrophy in Wolfram syndrome causes bilateral central visual field loss, leading to legal blindness in the majority of patients^44–46^. Annual ophthalmological assessment is recommended for all patients and should include best-corrected visual acuity (with cycloplegic refraction in children), color vision, intraocular pressures, slit-lamp examination of the anterior and posterior segments, visual fields (if age-appropriate), and optical coherence tomography (OCT) of the optic nerve and macula (if age-appropriate). Subclinical retinal nerve fiber layer thinning may precede symptomatic visual loss and is best detected with OCT imaging; macular ganglion cell layer measurements on OCT may precede retinal nerve fiber layer changes.

***Statement 7 There is no proven treatment that can slow the progression of visual loss in Wolfram Spectrum Disorders. Management remains supportive, including referral to a low-vision clinic where available, registration as severely sight impaired (if applicable in country), occupational rehabilitation, and special educational needs support (consensus* ≥*80%; expert opinion; Round 2)*.**

There is currently no proven treatment to slow the progression of visual loss in Wolfram Spectrum Disorders. Management therefore remains supportive. Referral to low vision services should be made promptly when functional impairment arises. Depending on national systems, patients should be registered as severely sight impaired, referred for occupational rehabilitation, and assessed for special educational needs as appropriate. Certification of sight loss, where available, can provide access to additional services and concessions that may benefit the patient.

### Neurology

***Statement 8 At their annual neurology review (or every 2 years if stable), all patients should be assessed for mental status and cognition (2-step commands, serial 7s, reading, repetition; MoCA/Blind-MoCA or neuropsychology evaluation if abnormal); cranial nerve function (smell, taste, dysarthria, facial symmetry, swallowing or choking difficulties, apneic episodes); motor function (tone, power); sensory function (vibration, temperature, pin-prick); reflexes; coordination; and gait. Speech-language therapy, occupational therapy, and physiotherapy should be considered (consensus* ≥*80%; expert opinion; Round 2)*.**

Progressive neurodegeneration is the hallmark of later-stage disease and the primary determinant of mortality, and assessment should begin with clinical examination before neuroimaging. Magnetic resonance imaging (MRI) demonstrates progressive atrophy of the brainstem, cerebellum, thalamus, and basal ganglia^47–49^. Clinical manifestations include cerebellar ataxia, dysarthria, dysphagia, autonomic dysfunction, and ultimately respiratory failure^5^.

Neurological involvement in Wolfram syndrome is progressive, diverse, and frequently underrecognized. Annual neurological review (or every two years if clinically stable) should include systematic assessment of cognition, cranial nerve function, motor function, sensory function, coordination, reflexes, and gait. Speech-language therapy, occupational therapy, and physiotherapy should be considered at each review.

***Statement 9 Patients with imbalance should be referred to a physiotherapist or physical therapist to assess gait, fall-risk, and learn balance exercises. If vestibular dysfunction is suspected, refer to ENT, occupational therapy, and/or audiology (consensus* ≥*80%; expert opinion; Round 2)*.**

Specific neurological complications require targeted management. Patients with balance impairment should be referred to physiotherapy for gait assessment, fall-risk stratification, and exercise prescription. Vestibular dysfunction warrants ENT referral. Autonomic dysfunction, evidenced by orthostatic hypotension and temperature dysregulation, should be assessed annually by measurement of lying and standing blood pressure. Chronic fatigue should be systematically assessed using a validated questionnaire, such as the Fatigue Severity Scale^50^, at each clinic visit.

***Statement 10 Patients diagnosed early in life should be assessed for hearing loss every 0.5–1 year for the first 4–5 years of life: auditory brainstem responses (or evoked otoacoustic emissions if unavailable); assessment for hearing aids; audiogram, speech discrimination tests, and tympanometry (age-dependent). After the first 5 years of life, review hearing at a minimum of every 2 years, with earlier reassessment if acute deterioration, device change, or key school transition (consensus* ≥*80%; expert opinion / patient feedback; Round 3)*.**

Sensorineural deafness is present in approximately 60–70% of patients, predominantly affecting high frequencies^51–53^. Assessment and appropriate amplification are important determinants of speech development and quality of life.

Hearing assessment is critical, particularly in young children, as early intervention with hearing amplification is an important determinant of speech and language development; cochlear implantation is generally reserved for patients with profound early-onset (autosomal dominant) deafness rather than for childhood-or adult-onset Wolfram syndrome. For patients diagnosed in infancy, formal audiological testing (rather than caregiver report alone) every 6–12 months for the first 4–5 years of life is recommended, with biennial review thereafter once hearing is stable. Swallowing assessment by a speech and language specialist is indicated if dysphagia, dysarthria, weight loss, recurrent chest infections, or prolonged mealtimes are reported.

***Statement 11 If there are concerns for dysphagia, dysarthria, or features suggestive of swallowing difficulties (weight loss, recurrent chest infections, choking, prolonged mealtimes), patients should be assessed by a speech and language specialist, gastroenterologist, or other relevant specialist per local standard of care (consensus*** ≥***80%; expert opinion; Round 2)*.**

Swallowing difficulties may reflect progressive brainstem and cranial nerve involvement and carry a risk of aspiration and malnutrition ^5,54^. Where dysphagia, dysarthria, weight loss, recurrent chest infections, choking, or prolonged mealtimes are reported, prompt assessment by a speech and language specialist, gastroenterologist, or other relevant specialist should be arranged according to local standards of care^55^.

***Statement 12 If symptoms or signs suggestive of sleep apnea are present (morning headaches, snoring, excessive daytime sleepiness), patients should be assessed with polysomnography (consensus*** ≥***80%; expert opinion; Round 1)*.**

Sleep-disordered breathing, including central and obstructive sleep apnea, may arise from brainstem involvement and contribute to fatigue, morning headaches, and daytime sleepiness.

Where suggestive symptoms or signs are present, polysomnography should be undertaken and treatment instituted as indicated ^56^.

***Statement 13 Patients may experience autonomic dysfunction including temperature dysregulation and fluctuating blood pressure. Orthostatic vitals should be checked at least annually: heart rate and blood pressure should be measured after 5 minutes supine, then after standing for 1 minute and 3 minutes (consensus* ≥*80%; expert opinion / patient feedback; Round 3)*.**

Autonomic dysfunction is common and may manifest as orthostatic intolerance, temperature dysregulation, and fluctuating blood pressure. Orthostatic vital signs should be checked at least annually, with heart rate and blood pressure measured after 5 minutes supine and again after standing for 1 and 3 minutes, and symptomatic postural hypotension managed with conservative measures in the first instance ^57^.

***Statement 14 Chronic fatigue is common and should ideally be assessed with a validated questionnaire (e.g., Fatigue Severity Scale) at clinic appointments. Management includes screening for and treating underlying causes (anxiety, depression, anemia, sleep disorders, thyroid disease), patient information, sleep hygiene, hydration and nutrition, and supervised graded aerobic exercise. Referral to physiotherapy or occupational therapy should be considered where available (consensus* ≥*80%; expert opinion / patient feedback; Round 3)*.**

Chronic fatigue is a frequent and disabling symptom that significantly affects quality of life. Assessment with a validated tool such as the Fatigue Severity Scale helps quantify severity and monitor change ^50^. Management should include screening for and treating contributory factors such as anxiety, depression, anemia, sleep disorders, and thyroid disease, alongside advice on sleep hygiene, hydration, nutrition, and supervised graded aerobic exercise, with referral to physiotherapy or occupational therapy where available.

***Statement 15 Migraines and headaches are common in Wolfram syndrome and can be exacerbated by systemic infections. Reversible causes should be screened for and treated (dehydration, infection, poor glycemic control, poor sleep hygiene). Acute and preventative management depends on headache type, severity, and age (consensus* ≥*80%; expert opinion; Round 2)*.**

Headaches and migraines are common in Wolfram syndrome and may be exacerbated by systemic infections, dehydration, poor glycemic control, and poor sleep^58,59^. Reversible triggers should be identified and addressed before escalating to specialist headache management. Brain MRI without contrast, with thin cuts through the brainstem and orbits, should be obtained at diagnosis and when new or worsening neurological impairment is present. Movement disorders including ataxia, tremors, and parkinsonism may be manifestations of Wolfram syndrome-related neurodegeneration but may also reflect co-existing pathology and should prompt neurological review.

***Statement 16 MRI brain without contrast, with thin cuts through the brainstem and orbits, should be considered in patients with a new diagnosis of Wolfram syndrome or with new or worsening neurological impairment. Wolfram syndrome patients show progressive atrophy of the cerebellum, brainstem, thalamus, and/or basal ganglia (consensus* ≥*80%; expert opinion; Round 3)*.**

Brain MRI without contrast, with thin cuts through the brainstem and orbits, supports diagnosis and characterization of neurodegeneration. Imaging typically demonstrates progressive atrophy of the cerebellum, brainstem, thalamus, and basal ganglia ^47,49^. MRI should be considered at new diagnosis and when new or worsening neurological impairment is identified.

***Statement 17 Movement disorders (ataxia, tremors, myoclonus, parkinsonism, tics) may be present in Wolfram syndrome, but other underlying diagnoses may co-exist and should be considered as necessary (consensus*** ≥***80%; expert opinion / patient feedback; Round 3)*.**

Movement disorders, including ataxia, tremor, myoclonus, parkinsonism, and tics, may occur as part of Wolfram syndrome-related neurodegeneration. Because co-existing or alternative diagnoses can produce similar features, these presentations should prompt neurological review and consideration of other underlying causes where clinically appropriate^5,60^.

***Statement 18 Seizures are not common in Wolfram syndrome. If they occur, investigations should be undertaken to establish the underlying cause (consensus* ≥*80%; expert opinion; Round 2)*.**

Seizures are uncommon in Wolfram syndrome. When they do occur, they should not be assumed to be a direct manifestation of the underlying disorder, and appropriate investigation should be undertaken to establish and treat the underlying cause^61^.

### Endocrinology

***Statement 19 To diagnose diabetes mellitus in Wolfram syndrome, follow ISPAD clinical criteria: (1) classic symptoms or hyperglycemic crisis with plasma glucose* ≥*11.1 mmol/L (200 mg/dL); or (2) fasting plasma glucose* ≥*7.0 mmol/L (125 mg/dL); or (3) 2-hour post-load glucose* ≥*11.1 mmol/L during OGTT (200 mg/dL); or (4) HbA1c* ≥*48 mmol/mol (6.5%). In the absence of unequivocal hyperglycemia, two abnormal test results are required. The absence of islet cell antibodies at diagnosis raises suspicion that diabetes mellitus is not type 1 (consensus* ≥*80%; level B; Round 2)*.**

Diabetes mellitus is the presenting feature in most patients, typically appearing in the first decade of life (median onset 6 years)^2,6,34,62^. It is characterized by insulin deficiency, the absence of islet autoantibodies, and sometimes residual C-peptide more than a year after diagnosis, features that distinguish it from type 1 autoimmune diabetes and are critical for correct diagnosis.

Diabetes mellitus in Wolfram syndrome is insulin-deficient but distinct from type 1 autoimmune diabetes. Diagnosis follows established International Society for Pediatric and Adolescent Diabetes (ISPAD) criteria^63^. Critically, the absence of islet autoantibodies at diagnosis should prompt consideration of Wolfram syndrome and genetic testing in any child with unexplained insulin-requiring diabetes.

***Statement 20 Where feasible, continuous glucose monitoring as part of a hybrid closed-loop insulin delivery system is the preferred method for managing diabetes mellitus in Wolfram syndrome. Hypoglycemia alarms should ideally be both audible and vibrating, with adaptation for vision loss if required (consensus* ≥*80%; level C; Round 2)*.**

Insulin is the mainstay and cornerstone of treatment for diabetes mellitus in Wolfram syndrome. Management of diabetes mellitus should prioritize continuous glucose monitoring integrated with hybrid closed-loop insulin delivery where available^64,65^. Hypoglycemia alarm systems should incorporate both audible and vibrating alerts, with adaptations for visual impairment.

***Statement 21 Children with Wolfram syndrome–related diabetes mellitus should have glycemic control monitored every 3 months (adults every 6 months), using HbA1c, insulin dosing, and continuous glucose monitoring (if available). Pre-diabetic patients should have HbA1c assessed every 4 months. Patients should be taught to seek medical assistance promptly if glucose levels are not well controlled (consensus* ≥*80%; level B / expert opinion; Round 3)*.**

Glycemic control in Wolfram syndrome-related diabetes mellitus requires regular structured review, incorporating HbA1c, insulin dosing review, and continuous glucose monitoring data where available.

The recommended intervals of every 3 months in children and every 6 months in adults represent minimum review frequencies for clinically stable patients who are meeting their glycemic targets, consistent with the American Diabetes Association Standards of Care^66^. These intervals are intended as a floor rather than a ceiling. Review should be intensified to at least every 3 months for adults who are not meeting glycemic goals, who have had recent treatment changes, or who experience frequent or severe hypoglycemia or hyperglycemia or other changes in health status, in line with the same guidance.

The 4-month interval recommended for genetically diagnosed individuals who have not yet developed diabetes reflects a different objective than the intervals used for established diabetes. Rather than glycemic management, its purpose is surveillance for conversion, allowing early detection of diabetes onset in patients carrying a confirmed genetic diagnosis of Wolfram syndrome. This approach parallels the established framework for monitoring presymptomatic individuals at risk of diabetes, in which periodic assessment of glycemic status by fasting glucose, oral glucose tolerance testing, and HbA1c is performed at intervals that shorten with higher risk of progression ^67,68^. Because Wolfram syndrome diabetes is non-autoimmune, surveillance relies on these glucose-based measures rather than islet autoantibodies^2^.

The two sets of intervals therefore serve distinct purposes, surveillance for conversion versus management of established disease, and do not represent a hierarchy of monitoring intensity. Patients and families should be educated to recognize early signs of dysglycemia and to seek medical assistance promptly when glucose levels are not well controlled^2^.

***Statement 22 Arginine vasopressin deficiency (formerly diabetes insipidus) should be diagnosed using paired early morning urine and fasting plasma osmolality and sodium measurements after nocturnal and morning euglycemia. A water deprivation test is a second-line test and should only be undertaken in a specialist center with appropriate supervision (consensus* ≥*80%; level B / expert opinion; Round 1)*.**

Arginine vasopressin deficiency (formerly known as central diabetes insipidus) results from progressive loss of hypothalamic vasopressin-producing neurons and affects the majority of patients^5^. It contributes substantially to morbidity through polyuria, polydipsia, and risk of hypernatremic dehydration.

Arginine vasopressin deficiency (formerly central diabetes insipidus) should be diagnosed by measuring paired early-morning urine and plasma osmolality in the setting of euglycemia.

Additional neuroendocrine complications, including growth failure, adrenocortical insufficiency, pubertal delay, hypogonadism, and hypothyroidism, should be screened for with targeted investigations as clinically indicated ^69^.

***Statement 23 First-line treatment for arginine vasopressin deficiency in Wolfram syndrome is usually oral, sublingual, or intranasal desmopressin (consensus*** ≥***80%; level B / expert opinion; Round 1)*.**

Desmopressin, administered orally, sublingually, or intranasally, is the first-line treatment for arginine vasopressin deficiency. Dosing should be individualized and monitored to avoid hyponatremia, with particular attention to fluid balance during intercurrent illness.

***Statement 24 If there are clinical signs of growth failure, adrenocortical insufficiency, pubertal delay, or underactive thyroid, referral to an endocrinologist for anterior pituitary function tests should be considered (consensus*** ≥***80%; expert opinion; Round 1)*.**

Anterior pituitary dysfunction may accompany posterior pituitary involvement. Where there are clinical signs of growth failure, adrenal insufficiency, pubertal delay, or hypothyroidism, referral to an endocrinologist for anterior pituitary function testing should be considered so that hormone deficiencies can be identified and treated^70^.

***Statement 25 To screen for hypogonadism in Wolfram syndrome, take an age-directed history, assess Tanner stage, and measure serum testosterone or estradiol. Second-line tests include FSH, LH, inhibin B, anti-Müllerian hormone, sperm count (males), and dynamic function tests. Management of hypogonadism includes hormone replacement therapy directed by an endocrinologist (consensus* ≥*80%; expert opinion; Round 1)*.**

Hypogonadism may occur and warrants assessment through an age-directed history, Tanner staging, and measurement of serum testosterone or estradiol. Second-line investigations include FSH, LH, inhibin B, anti-Mullerian hormone, sperm count in males, and dynamic function tests. Hormone replacement therapy, where indicated, should be directed by an endocrinologist ^71^.

***Statement 26 Fertility counseling and preservation should be offered in a timely manner if subfertility or infertility is suspected. There are case reports of successful pregnancies and of affected males who have fathered children (consensus* ≥*80%; level C / expert opinion; Round 1)*.**

Subfertility and infertility may affect some individuals, although successful pregnancies and affected males who have fathered children have been reported. Fertility counseling and preservation should be offered in a timely manner where subfertility or infertility is suspected, ideally before fertility is further compromised ^72,73^.

***Statement 27 Height and weight should be monitored in all patients by plotting on standard growth charts. Height should be measured in adults to calculate BMI (consensus* ≥*80%; expert opinion; Round 2)*.**

Growth and nutritional status should be monitored in all patients by plotting height and weight on standard growth charts, with height measured in adults to calculate body mass index.

Deviation from expected growth trajectories should prompt assessment for endocrine and nutritional contributors^74–77^.

***Statement 28 Patients should be screened for dyslipidemia annually if diabetes mellitus is present (consensus* ≥*80%; expert opinion; Round 1)*.**

Patients with diabetes mellitus are at increased cardiometabolic risk, and dyslipidemia is an important modifiable contributor. Annual lipid screening is recommended where diabetes mellitus is present, with management according to established diabetes care standards ^78^.

### Urology and gastroenterology

***Statement 29 Baseline urological assessment at diagnosis should include a standardized questionnaire regarding urinary symptoms and a voiding diary; clinical examination and assessment of renal function; blood electrolytes, urea, creatinine, and GFR; bladder and renal ultrasound; and non-invasive urodynamic testing (consensus*** ≥***80%; expert opinion; Round 2)*.**

Neurogenic bladder resulting from autonomic neuropathy affects the majority of patients with advancing disease and carries significant risks of upper urinary tract damage and renal impairment if not identified and managed early.

Neurogenic bladder resulting from autonomic neuropathy is a major source of morbidity in Wolfram syndrome^79,80^. Urological assessment should commence at diagnosis and include a urinary symptoms questionnaire, voiding diary, clinical examination, serum electrolytes, urea, creatinine, and glomerular filtration rate, a urine albumin-to-creatinine ratio, blood pressure measurement, bladder and renal ultrasound, and non-invasive urodynamic testing. Follow-up urological assessment should occur every 1–2 years or earlier if symptomatic. Clinicians should be aware of acute bladder outflow tract obstruction causing acute kidney injury; there is also an increased risk of urinary tract infections due to urinary stasis.

***Statement 30 Within the first year of diagnosis, baseline screening for neurogenic bladder should occur with non-invasive urodynamic function testing, validated questionnaires, and renal ultrasound where indicated. Follow-up screening should occur every 1–2 years (or sooner if symptomatic), at the discretion of the urologist or medical care provider (consensus* ≥*80%; expert opinion; Round 3)*.**

Baseline screening for neurogenic bladder should be undertaken within the first year of diagnosis using non-invasive urodynamic function testing, validated questionnaires, and renal ultrasound where indicated, complementing the broader baseline urological assessment outlined above by specifying the timing and the recommended follow-up schedule. Follow-up screening every 1-2 years, or sooner if symptomatic, allows early detection of bladder dysfunction before irreversible upper urinary tract damage occurs; the aetiology may relate to a polyuric stretch injury or to autonomic nerve dysfunction ^79^.

**Statement 31 *Treatment options for neurogenic bladder in Wolfram syndrome should be discussed in conjunction with a pediatric or adult urology team (consensus* ≥*80%; expert opinion; Round 1)*.**

Treatment of neurogenic bladder should be planned in conjunction with a pediatric or adult urology team. Management depends on whether the bladder is overactive or underactive and typically begins with bladder training, including double voiding, and oral therapeutics; desmopressin may reduce urine output and improve continence. With progression to megacystis and refractory incontinence, clean intermittent self-catheterization is often required, with later options including an indwelling catheter or a Mitrofanoff appendicovesicostomy ^79^.

***Statement 32 First-line management of fecal incontinence should include evaluating for and managing constipation-related encopresis or overflow soiling through diet optimization (adequate fiber and water intake), recommending a bowel regimen, and considering pelvic floor physical therapy where possible (consensus*** ≥***80%; expert opinion; Round 3)*.**

Gastrointestinal manifestations including dysmotility, esophageal spasms, and constipation are recognized features of Wolfram syndrome attributable to autonomic neuropathy. Fecal incontinence and bowel dysfunction are increasingly recognized manifestations, arising from autonomic neuropathy affecting the enteric nervous system. Bowel dysfunction in Wolfram syndrome spans a spectrum from constipation and intestinal dysmotility to intractable fecal incontinence, and has a severe impact on quality of life ^81^. Fecal incontinence warrants active clinical inquiry at each review.

Bowel dysfunction, including constipation, intestinal dysmotility, and fecal incontinence, is an underrecognized but clinically important manifestation of Wolfram syndrome attributable to autonomic neuropathy^81,82^. The spectrum ranges from constipation and intestinal pseudo-obstruction to intractable fecal incontinence, and bowel dysfunction has a severe impact on quality of life. Fecal incontinence, when present, should be evaluated for constipation-related overflow soiling before assuming a primary neurogenic cause. First-line management includes dietary optimization (adequate fiber and fluid intake), a structured bowel regimen, and pelvic floor physical therapy where accessible. Where neurogenic fecal incontinence is confirmed or suspected, specialist gastroenterological assessment should be considered.

***Statement 33 Management options for constipation in Wolfram syndrome include diet and behavior advice, stool softeners, fiber supplements, laxatives, and disimpaction regimens (consensus*** ≥***80%; expert opinion; Round 1)*.**

Constipation is common and may reflect autonomic neuropathy affecting the enteric nervous system. Management options include dietary and behavioral advice, stool softeners, fiber supplements, laxatives, and disimpaction regimens, escalated in a stepwise fashion and coordinated with specialist input where symptoms are refractory ^83^.

### Psychiatry and cognition

***Statement 34 All newly diagnosed patients should be offered assessment by an appropriate healthcare professional (psychologist or psychiatrist) to identify emotional, developmental, and educational needs and to signpost to support (consensus*** ≥***80%; expert opinion; Round 3)*.**

Psychiatric illness, particularly depression, anxiety, and hallucinations, is underrecognized in Wolfram syndrome and contributes substantially to patient burden. Neuropsychiatric conditions including attention-deficit/hyperactivity disorder (ADHD) and obsessive-compulsive disorder (OCD)-related symptoms are recognized and should be assessed as part of routine psychiatric review. Proactive psychiatric assessment and treatment are critical components of care.

Psychiatric illness is one of the most significant and underappreciated sources of morbidity in Wolfram syndrome^84,85^. Depression, anxiety, and psychosis affect a substantial proportion of patients, and psychiatric symptoms may precede or accompany neurological deterioration. All newly diagnosed patients should be offered assessment by a psychologist or psychiatrist to identify emotional, developmental, and educational needs, and to facilitate access to appropriate support services.

***Statement 35 First-line treatment for anxiety and depression in Wolfram syndrome should be psychotherapy and SSRIs, managed in consultation with a psychiatrist. There is limited evidence that fluoxetine and citalopram may be more effective than sertraline; however, other SSRIs can be used as clinically indicated (consensus* ≥*80%; level C / expert opinion; Round 3)*.**

First-line pharmacological treatment for anxiety and depression is selective serotonin reuptake inhibitor (SSRI) therapy, managed in consultation with a psychiatrist. Limited case series data suggest that fluoxetine and citalopram may be particularly effective in Wolfram syndrome, though other SSRIs remain appropriate when clinically indicated ^85^. Psychotherapy should be offered alongside pharmacological treatment where available.

## Discussion

These guidelines represent the first international, multidisciplinary, patient-inclusive clinical consensus for Wolfram syndrome and establish a common framework for diagnosis, monitoring, and management across all major organ systems. They are intended to improve the consistency and quality of clinical care worldwide, including in settings far from specialist centers, where most patients receive their care.

The methodological approach, a three-round international Delphi process informed by a systematic evidence review and incorporating formal patient and family feedback, reflects established best practice in rare disease guideline development.^32^ The achievement of ≥80% consensus on all 35 final statements across 65 international specialists spanning five continents and six clinical disciplines provides a strong foundation of expert agreement. The parallel patient consultation, formally integrated through international advocacy organizations, represents an advance over prior rare disease guideline processes and ensures that the perspectives of those living with Wolfram syndrome are embedded in the recommendations.

Several clinical messages emerge from these guidelines. First, early and accurate genetic diagnosis is essential: Wolfram syndrome is systematically underdiagnosed, and the absence of islet autoantibodies in a child with apparent type 1 diabetes should prompt genetic testing, ideally on a large gene panel so that alternative monogenic causes of diabetes can also be detected, rather than targeted sequencing of *WFS1* alone. Second, multidisciplinary coordinated care is mandatory. No single specialty can manage the full clinical complexity of this disease. Third, proactive management of preventable complications, particularly neurogenic bladder, psychiatric illness, and autonomic dysfunction, can substantially reduce morbidity. Fourth, patient advocacy organizations are integral partners in care, not adjuncts.

Palliative and supportive care also merit explicit consideration. Because Wolfram syndrome is progressive and, in its advanced stages, associated with substantial neurological disability and reduced life expectancy, early integration of palliative care can improve quality of life for patients and their families. Conversations about goals of care and advance care planning should be introduced sensitively, revisited as the disease evolves, and embedded within routine multidisciplinary review rather than reserved for times of crisis. Access to symptom management, psychological support, and, where appropriate, hospice services should be considered alongside disease-directed treatment^71^.

Important limitations should be acknowledged. The evidence base for Wolfram syndrome remains predominantly observational, as there are no randomized controlled trials to guide practice, and most recommendations therefore rest on expert consensus. This reflects both the global rarity of the disease and the challenges of securing research funding for rare conditions. Future iterations of these guidelines should incorporate prospective audit data and, where available, clinical trial evidence, particularly from ongoing therapeutic trials targeting ER stress pathways in Wolfram syndrome.

### Future directions

Several disease-modifying strategies are now under active investigation in Wolfram syndrome. Alongside approaches aimed directly at the underlying endoplasmic reticulum stress and mitochondrial dysfunction, repurposing of established diabetes therapies has attracted particular interest^86–88^.

Limited evidence from preclinical studies and a small number of case reports suggests that glucagon-like peptide-1 (GLP-1) receptor agonists may be beneficial in Wolfram syndrome. Reported effects include reduced insulin requirements and improved glycemic control, with preclinical data raising the possibility that these agents could slow disease progression. A recent case series from Aotearoa, New Zealand describes the phenotype and treatment response in affected individuals^89–94^. In preclinical models, GLP-1 receptor agonist therapy has been reported to ameliorate visual neurodegeneration and sensorineural hearing loss, raising the possibility of benefits beyond glycemic control, although this has not been demonstrated in patients^92,95^. These agents are currently one possible adjunctive option rather than a replacement for insulin, and should not completely replace insulin in children. Their known gastrointestinal side effects must be carefully considered in the context of existing autonomic dysfunction and growth delay in some patients.

These guidelines provide a comprehensive framework for improving the diagnosis and clinical management of Wolfram syndrome internationally. Coordinated multidisciplinary care, early genetic diagnosis, and proactive management of evolving complications are the pillars of good clinical practice for this disease. Implementation of these recommendations should be accompanied by a prospective audit to expand the evidence base and support future updates.

With a growing pipeline of therapeutic candidates targeting ER stress and mitochondrial dysfunction in Wolfram syndrome, there is an increasingly urgent need to establish international networks that can rapidly develop and validate the outcomes that matter most to patients, deliver consistent best supportive care today, and provide high-quality infrastructure for future randomized controlled trials.

## International Wolfram Syndrome Clinical Guidelines Consortium - Members

The following medical and allied health professionals with experience in the clinical care of individuals with Wolfram syndrome contributed to the modified three-round Delphi process by providing feedback, suggestions, and evaluations that helped refine the statements and inform these consensus recommendations.

**Fumihiko Urano**, Washington University, St. Louis, USA

**Timothy Barrett**, Birmingham Children’s Hospital and University of Birmingham, UK

**Saumel Ahmadi**, Washington University, St. Louis, USA

**Patrick Yu-Wai-Man**, University of Cambridge, UK

**Sarah Gladstone**, The Snow Foundation, St Louis, USA

**Josephine Elliott**, University of Birmingham, UK

**Bess Marshall**, Washington University, St Louis, USA

**Alanna Strong**, Children’s Hospital of Philadelphia, USA

**Wendy K. Chung**, Boston Children’s Hospital and Harvard Medical School, Boston, USA

**Angela M. Reiersen**, Washington University, St Louis, USA

**Tamara Hershey**, Washington University, St Louis, USA

**Paul F. Austin**, Texas Children’s Hospital and Baylor College of Medicine, USA

**Douglas Coplen**, St Louis Children’s Hospital and Washington University, St Louis, USA

**Christina Gurnett**, Children’s Hospital of Philadelphia and University of Pennsylvania, USA

**Kyle O. Rove**, University of Colorado and Children’s Hospital Colorado, USA

**Natasha Leibel**, Columbia University, New York, USA

**Yunshuo Tang**, Washington University, St Louis, USA

**Louis Philipson**, University of Chicago, Chicago, USA

**Rudolph L. Leibel**, Columbia University, New York, USA

**Irl B. Hirsch**, University of Washington, Seattle, USA

**Gregory Van Stavern**, Washington University, St Louis, USA

**Christophe Orssaud**, Hôpital Européen Georges Pompidou, Paris, France

**Renuka P. Dias**, Birmingham Children’s Hospital and University of Birmingham, UK

**Raniero Chimienti**, Vita-Salute San Raffaele University, Milan, Italy

**Agathe Roubertie**, Centre Hospitalier Universitaire de Montpellier, Montpellier, France

**Ben Wright**, Queen Elizabeth Hospital, Birmingham, UK

**Arunkumar R. Pande**, Lucknow Endocrine Diabetes and Thyroid Clinic and Health City Vistaar, Lucknow, India

**Gema Esteban-Bueno**, Clinical Management Unit Almería Periférica, Andalusian Health Service, Almería, Spain; Spanish Association for Research and Help to Wolfram Syndrome, Almería, Spain

**Kalpana Suhas Jog**, King Edward Memorial Hospital and Research Centre, Pune, India

**Susan Gleeson**, Birmingham Children’s Hospital, UK

**Mariam Oniani**, M. Iashvili Children’s Central Hospital, Georgia

**Paulina Cruz Bravo**, Washington University, St Louis, USA

**Amy Robichaux-Viehoever**, Washington University, St Louis, USA

**James Hoekel**, Washington University, St Louis, USA

**Leanne Stunkel**, Washington University, St Louis, USA

**Lauren Golden**, New York University, New York, USA

**Carolina Fischinger Moura de Souza**, Casa dos Raros, Porto Alegre, Brazil

**Anne Rochtus**, University Hospital Leuven, Belgium

**Esko Wiltshire**, University of Otago Wellington and Health New Zealand Capital, Coast and Hutt Valley, New Zealand

**Giulio Frontino**, Department of Pediatrics, IRCCS Ospedale San Raffaele, Milano, Italy

**Amelia Caretto**, Diabetology and Nutrition Unit, ASST Santi Paolo e Carlo, Milan, Italy

**Denise Williams**, NHS England, UK

**Archana Kulkarni**, Birmingham Women’s and Children’s Hospital, UK

**Agnieszka Zmysłowska**, Medical University of Lodz, Poland

**Sam Peter Gurney**, Birmingham Children’s Hospital, UK

**Rinki Murphy**, University of Auckland and Auckland Diabetes Centre, Auckland, New Zealand

**Sofia Salahuddin**, Queen Elizabeth University Hospital, Birmingham, UK

**Alfredo A. Sadun**, Doheny Eye Institute, University of California, Los Angeles, USA

**Marie McGee**, Birmingham Women’s and Children’s NHS Trust, UK

**Kristin A. Maloney**, University of Maryland School of Medicine, Baltimore, USA

**Stephen I. Stone**, Washington University, St Louis, USA

**Yukio Tanizawa**, Yamaguchi University, Japan

**Stacy Hurst,** Washington University, St. Louis, USA

**Ashley Raterman,** Washington University, St. Louis, USA

**Marcela Votruba**, Cardiff University and University Hospital Wales, UK

**Leonardo Caporali**, IRCCS Istituto delle Scienze Neurologiche di Bologna, Bologna, Italy

**Claudio Fiorini**, IRCCS Istituto delle Scienze Neurologiche di Bologna, Bologna, Italy

**Raoul Kanav Khanna**, Bretonneau University Hospital and INSERM iBraiN U1253, Tours, France; University of Cambridge and Cambridge University Hospitals, Cambridge, UK

**Chiara La Morgia**, IRCCS Istituto delle Scienze Neurologiche di Bologna, Bologna, Italy; University of Bologna, Italy

**Susan Patricia Mollan**, Queen’s University, Kingston, Ontario, Canada

**An-Guor Wang**, Taipei Veterans General Hospital, Taiwan

**Piero Barboni**, San Raffaele University, Milan, Italy

**Alina Radu**, Hôpital Européen Georges Pompidou and Université Paris Cité/Assistance Publique-Hôpitaux de Paris, Paris, France

**Jacques Beltrand,** Hôpital Necker-Enfants Malades and Université Paris Cité/Assistance Publique-Hôpitaux de Paris, Paris, France

**Neringa Jurkute**, Moorfields Eye Hospital NHS Foundation Trust, London, UK; UCL Institute of Ophthalmology, London, UK; The National Hospital for Neurology and Neurosurgery, UCL Queen Square Institute of Neurology, London, UK

**Emilio Hideyuki Moriguchi**, Universidade Federal do Rio Grande do Sul, Brazil

**Elmarie van der Merwe**, Birmingham Children’s Hospital, UK

**Julia Rohayem**, Ostschweizer Kinderspital St. Gallen, Switzerland

**Mars Skae**, Royal Manchester Children’s Hospital, UK

**Andrew R. Lee**, Washington University, St Louis, USA

**Andrew Hattersley**, University of Exeter, UK

**Maggie Shepherd**, Royal Devon University Healthcare NHS Foundation Trust and University of Exeter, UK

**Kashyap Patel**, University of Exeter, UK

**Kevin Colclough**, University of Exeter, UK

**Valerio Carelli**, IRCCS Istituto delle Scienze Neurologiche di Bologna, Bologna, Italy; University of Bologna, Italy

**Michele Carbonelli**, University of Bologna, Italy

**Orit Pinhas-Hamiel**, Pediatric Endocrine and Diabetes, Sheba Medical Center, Ramat Gan, Israel; Gray Faculty of Medical and Health Sciences, Tel Aviv University, Tel Aviv, Israel

**Noga Minsky**, Division of Endocrinology, Diabetes and Metabolism, Sheba Medical Center, Ramat Gan, Israel; Gray Faculty of Medical and Health Sciences, Tel Aviv University, Tel Aviv, Israel

**Odelia Chorin**, The Danek Gertner Institute of Human Genetics, Sheba Medical Center, Ramat Gan, Israel; Gray Faculty of Medical and Health Sciences, Tel Aviv University, Tel Aviv, Israel

**Gil Leibowitz**, Endocrinology and Metabolism Service, Hadassah Medical Center, Jerusalem, Israel; Faculty of Medicine, The Hebrew University of Jerusalem, Jerusalem, Israel

**Avivit Cahn**, Internal Medicine Department A, Hadassah Hebrew University Hospital, Jerusalem, Israel; The Faculty of Medicine, Hebrew University of Jerusalem, Jerusalem, Israel

**Stephanie Gebel,** The Snow Foundation, St. Louis, USA

**Tracy Lynch,** Wolfram syndrome UK, UK

**Nolwen Le Floch,** Association du Syndrome de Wolfram, France

## Data Availability

All data produced in the present study are available upon reasonable request to the authors.

## Acknowledgements

The authors express their sincere thanks to the participating patient support and advocacy organizations and to the patients and families whose input shaped these recommendations.

## Author Contributions

FU and TB conceived and co-chaired the study and designed the modified three-round Delphi methodology. FU, TB, JE, SA, PYWM, SG, SGe, and TL constituted the international steering committee and together conducted the systematic literature review, drafted the candidate consensus statements across the six clinical domains, and iteratively revised the statements between voting rounds. SG, SGe, and TL led patient and family engagement and coordinated the parallel patient and caregiver survey through the participating international advocacy organizations. The members of the International Wolfram Syndrome Clinical Guidelines Consortium contributed to the modified three-round Delphi process in several ways, including rating candidate statements, providing structured feedback and suggestions to refine the statements, and contributing to the writing, review, and editing of the manuscript. JE, SA, FU, and TB analyzed the Delphi data and wrote the original draft. All authors and all members of the International Wolfram Syndrome Clinical Guidelines Consortium critically reviewed the manuscript and approved the final version.

